# Population-specific facial traits and diagnosis accuracy of genetic and rare diseases in an admixed Colombian population

**DOI:** 10.1101/2022.11.25.22282761

**Authors:** Luis Miguel Echeverry, Estephania Candelo, Eidith Gómez, Paula Solís, Diana Ramírez, Diana Ortiz, Alejandro González, Xavier Sevillano, Juan Carlos Cuéllar, Harry Pachajoa, Neus Martínez-Abadías

## Abstract

Up to 40% of genetic and rare disorders (RD) present facial dysmorphologies. Visual assessment of facial gestalt is commonly used for clinical diagnosis, health management and treatment monitoring. Quantitative approaches to facial phenotypes are more objective and provide first diagnoses of RD with relatively high accuracy, but are mainly based on populations of European descent, disregarding the influence of population ancestry. Here we assessed the facial phenotypes associated to four genetic disorders in a Latino-American population from Colombia. We recorded the coordinates of 18 facial landmarks in 2D images from 79 controls 51 pediatric individuals diagnosed with Down (DS), Morquio (MS), Noonan (NS) and Neurofibromatosis type 1 (NF1) syndromes. We quantified facial differences using Euclidean Distance Matrix Analysis (EDMA) and assessed the diagnostic accuracy of Face2gene, an automatic deep learning algorithm with widespread use in the clinical practice.

Quantitative comparisons indicated that individuals diagnosed with DS and MS were associated with the most severe phenotypes, with 58.2% and 65.4% of facial traits significantly different as compared to controls. The percentage decreased to 47.7% in NS and to 11.4% in NF1. Each syndrome presented a characteristic pattern of facial dysmorphology, supporting the potential of facial biomarkers for disorder diagnosis. However, our results detected population-specific traits in the Colombian population as compared to the facial gestalt described in literature for DS, NS and NF1. When clinical diagnosis based on genetic testing was used to verify the diagnosis based on 2D facial pictures, our results showed that Face2Gene accuracy was very high in DS, moderate in NS and NF1, and very low in MS, with low gestalt similarity scores in highly admixed individuals. Our study underscores the added value of precise quantitative comparison of facial dysmorphologies in genetic and rare disorders and the need to incorporate populations with diverse contributions of Amerindian, African and European ancestry components to further improve automatic diagnostic methods.

## INTRODUCTION

According to the Online Mendelian Inheritance in Man (OMIM) databank, there are more than 10,000 genetic and rare diseases (RD) affecting 7% of the world’s population (Nguengang Wakap et al., 2020; Viteri et al., 2020), which corresponds to approximately 500 million people. Although as a whole genetic and RD are a significant cause of morbidity and mortality in the pediatric population (Suárez-Obando, 2018), by separate each disorder affects a very reduced number of people. Depending on the country, the prevalence to consider a disease as rare ranges from 1 affected individual in 50,000 people to 1 in 200,000. This low prevalence has limited the research on rare disorders.

Currently, there is limited knowledge on the etiology of these disorders, a reduced percentage of diseases (20%) presents a known molecular basis associated to a detailed phenotype description, and treatment is only available for 0.04% of RD (Suárez-Obando, 2018). As orphan diseases, many RD are chronic and incurable, representing severe and debilitating conditions (Cortés, 2015). The diagnosis and management of genetic RD is currently a clinical challenge (Schieppati et al., 2008). Precise and early diagnosis is crucial for individuals and their families to get effective care and to reduce disease progression. However, due to the limited knowledge and complexity of these pathologies, diagnosis may take several years (Bannister et al., 2020) and people may experience a diagnostic odyssey with delays in their correct treatment and management (González-Lamuño & García Fuentes, 2008). For most rare diseases there are no reliable biomarkers for an early diagnosis (Gülbakan et al., 2016).

Among the wide constellation of clinical symptoms associated to genetic and rare disorders, craniofacial dysmorphologies emerge as potential biomarkers (Gurovich et al., 2019; Hallgrímsson et al., 2020). These phenotypes are highly prevalent (Bannister et al., 2020; Viteri et al., 2020) and are commonly used for diagnosis, management and treatment monitoring of genetic and RD (Bannister et al., 2020). Up to 40% of these disorders present characteristic craniofacial phenotypes, as in Down, Morquio, Noonan, Apert, Rett, Fragile X, Williams-Beuren and Treacher-Collins and velocardiofacial syndromes, as well as other conditions such as microcephaly, holoprosencephaly, palate/lip cleft and other 2,000 rare genetic disorders (Farrera et al., 2019; Hallgrímsson et al., 2020).

The genetic and environmental factors associated with these disorders alter the complex process that orchestrates facial morphogenesis during pre- and postnatal development, inducing facial dysmorphologies. Facial development is highly regulated by multiple signaling pathways (Martínez-Abadías et al., 2011; Richtsmeier & Flaherty, 2013; Hallgrímsson et al., 2015; Marcucio et al., 2015), including Fibroblast Growth Factor (*FGF*), Hedgehog (*HH*), Wingless (*WNT*) and Transforming Growth Factor Beta (*TGF-β*) and Bone Morphogenetic Proteins (*BPMs*). Disruptions in the regulation of any of these signaling pathways can lead to facial dysmorphogenesis (Kouskoura et al., 2011).

The facial patterns associated with each disorder are unique but vary within and among diagnostics, from subtle facial anomalies to severe malformations (Jones et al., 2021). In the clinical practice, craniofacial dysmorphology is commonly assessed through qualitative visual assessment and basic anthropometric measurements. However, this approach may not capture with maximum precision the anatomical complexity of the facial traits associated with these disorders. Qualitative descriptions of facial phenotypes are sometimes based on general terms such as coarse face, large and bulging head; saddle-like, flat bridged nose with broad, fleshy tip; or malformed teeth (Aase, 1990; Johannes et al., 2003; Reardon et al., 2007). Accurate identification of dysmorphic features for diagnosis depends on the clinician’s expertise and highly trained dysmorphologists are required to recognize the facial “gestalt” characteristic of many dysmorphic syndromes, especially for rarest disorders (Reardon et al., 2007).

Recent research seeks to incorporate into the clinical diagnosis of RD the use of objective and quantitative tools to assess facial phenotypes (Hammond et al., 2004; Hammond, 2007; Hammond and Suttie, 2012; Hurst, 2018; Köhler et al., 2019; Agbolade et al., 2020). Automated systems have been developed to improve and to accelerate the diagnostic process (Gurovich et al., 2019, Hallgrímsson et al., 2020, Hsieh et al., 2021). Within the clinical practice, Face2Gene is the most commonly used system (FDNA Inc., https://www.face2gene.com/), a community-driven phenotyping platform trained over 17,000 people representing more than 200 syndromes (Gurovich et al., 2019). Face recognition is performed on 2D images that can be collected with any type of digital camera or phone, without previous training. Syndrome classification is achieved using DeepGestalt, a cascade Deep Convolutional Neural Network (DCNN)-based method that achieved 91% top-10 accuracy in identifying the correct syndrome (Gurovich et al., 2019).

Diagnostic approaches based on 3D photogrammetry have been developed more recently (Hammond et al., 2004; Hammond, 2007; Hallgrímsson et al., 2020). The advantage of 3D facial models is that they are more efficient than 2D images in capturing the complexity of facial phenotypes, but their widespread use is limited because the photographic equipment required for generating 3D models is usually not commonly available in the clinical practice. Hallgrímsson et al. (2020) analyzed 3D facial models from 7,057 subjects including subjects with 396 different syndromes, relatives and unrelated unaffected subjects (https://www.facebase.org/). Deep phenotyping based on quantitative 3D facial imaging and machine learning presented a balanced accuracy of 73% for syndrome diagnosis (Hallgrímsson et al., 2020).

Automated methods have thus demonstrated high potential to facilitate the diagnosis of facial dysmorphic syndromes (Gurovich et al., 2019; Bannister et al., 2020; Hallgrímsson et al., 2020; Hsieh et al., 2021). These tools present high accuracy diagnosis in European and North American populations in which the software predictions are mostly based. However, the tools have not been thoroughly tested in populations with different ancestry, and it is not well understood the how facial phenotypes associated with genetic and RD might be influenced by the complex patterns of population ancestry characterizing human populations.

### Population ancestry in facial dysmorphologies: a long-disregarded factor

Facial shape shows wide variation across world-wide human populations (Xiong et al., 2019). Facial differences between populations detected in the shape of the forehead, brow ridges, eyes, nose, cheeks, mouth and jaw (Qiao et al., 2018) result from divergent evolutionary and adaptive histories of human populations occurred during the evolution of *Homo sapiens* over the last 200,000 years. Continuous migration and admixture keep shaping the facial phenotypes of human populations. Admixed populations display a variety of craniofacial morphologies, resembling the parental groups or presenting novel phenotypes (Martínez-Abadías et al., 2006), depending on dominance and epistatic interactions between alleles fixed or predominant in each parental group (Quinto-Sánchez et al., 2015). Therefore, the evolutionary and population dynamics of human populations results in genetic and phenotypic patterns that surrogate population ancestry (Ruiz-Linares et al., 2014; Sheehan and Nachman, 2014; Quinto-Sánchez et al., 2015) and can modulate the facial phenotypes associated to disease.

Few studies to date have analyzed the craniofacial phenotypes associated with genetic and RD in populations of non-European descent (Kruszka, et al., 2017; Kruszka, Addissie, et al., 2017; Kruszka, Sobering, et al., 2017; Dowsett et al., 2019), leaving African, Asian and Latin-American populations often disregarded and underrepresented. Thus, there is no reliable representation of facial phenotypes in genetic and rare diseases in populations of non-European descent. To develop quantitative approaches that efficiently diagnose genetic and RD in populations from all over the world, it is crucial to account for the influence of population ancestry on facial variation.

To contribute covering this gap, here we assessed the facial dysmorphologies associated to prevalent RD in a Latin-American population from the Southwest of Colombia. Latin-Americans are fascinating cases of hybrid/admixed populations that evolved on relatively short periods of time (Quinto-Sánchez et al., 2015; Mendoza-Revilla et al., 2021). Peopling of the Americas likely started 33,000 years ago (Ardelean et al., 2020; Becerra-Valdivia & Higham, 2020) by migration waves coming from North and South East Asia, and probably Australia (Castro E Silva et al., 2021). The first settlers arrived at North America and migrated towards southern regions following coastal and continental routes (González-José et al., 2003). Amerindian populations established all over the continent and adapted to a variety of environments over thousands of years. During the last 600 years, admixture with European and African populations has further shaped the genetic ancestry of Latin-American populations (Salzano & Bortolini, 2002; Salzano & Sans, 2014). In particular, the population from the region of Cali is the result of diverse migratory processes throughout the history of the Colombian settlement (Urrea-Giraldo & Álvarez, 2017). Admixture with the indigenous Amerindian population began in the sixteenth century with the arrival of the Spanish colonizers. In the eighteenth century, large colonial settlements of slaves brought from Africa were established in Cali for the exploitation of sugar cane and changed the population structure of Valle del Cauca. Nowadays, the population of Cali is characterized by indigenous and mestizo communities, with Amerindian and African ancestry components predominating over the European ancestry contribution (Urrea-Giraldo & Álvarez, 2017).

In this study, we compared the facial phenotypes associated to four genetic and RD, including Down syndrome, Mucopolysaccharidosis type IVA metabolic disorder known as Morquio syndrome, and two types of RASopathies, Noonan syndrome (NS) and neurofibromatosis type 1 (NF). The facial phenotype of these syndromes has not been previously characterized in Latin-American or Colombian populations and differences between populations with different ancestry backgrounds have not been assessed (Kruszka et al., 2017; Dowsett et al., 2019). Here, we quantitatively assessed the facial phenotypes associated to these syndromes and tested the hypothesis whether the diagnostic accuracy of automated methods commonly used in the clinical practice is affected by the admixed ancestry of this Colombian population of non-European descent.

## MATERIALS AND METHODS

### Participant recruitment for photographic sessions

The sample comprised 130 individuals from 0 to 18 years old from Valle del Cauca, a Southwest region in Colombia. The cohort included 79 age matched controls and 51 pediatric individuals diagnosed with Down, Morquio, Noonan and Neurofibromatosis type 1 syndromes that were recruited from the clinical genetics consultation at Hospital-Fundación Valle del Lili in Cali (Colombia), a tertiary health reference center for these genetic and rare disorders. See Table 1 for details on sample composition. In all participants, clinical diagnoses were confirmed by molecular genetic testing.

**Table 1.**
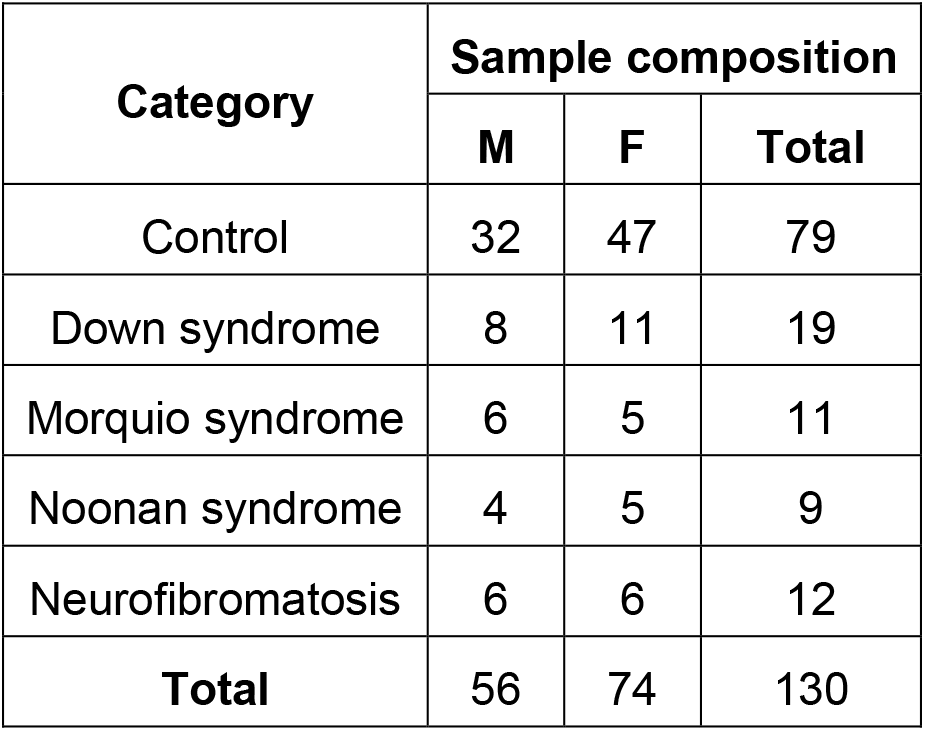
Human cohort by diagnostic group. Total number of participants as well as separated by sex (Male (M) / Female (F)) are provided.

| Category | Sample composition |  |  |
| --- | --- | --- | --- |
|  | M | F | Total |
| Control | 32 | 47 | 79 |
| Down syndrome | 8 | 11 | 19 |
| Morquio syndrome | 6 | 5 | 11 |
| Noonan syndrome | 4 | 5 | 9 |
| Neurofibromatosis | 6 | 6 | 12 |
| **Total** | 56 | 74 | 130 |

Down syndrome (DS, OMIM 190685) was first selected because it is one of the most common genetic disorders, and previous studies have shown that the clinical manifestations associated with DS vary across ethnicities (Kruszka et al., 2017). DS presents a worldwide prevalence of 14 per 10,000 live births, with life expectancy increasing from 25 to 60 years in developing countries (Glasson et al., 2002; Roper & Reeves, 2006; Patterson, 2009; Aivazidis et al., 2017). In most Latino-American regions, the real incidence of patients with DS remains unknown and is usually underreported. A cross-sectional study in Brazil reported a DS birth rate of 4 for every 10,000 live births (Laignier et al., 2021); whereas in Colombia several studies have reported a prevalence rate between 1 for every 1,000 to 5 per 10,000 live births (Hernández Ramírez & Manrique Hernández, 2006; Valencia Arana et al., 2008). DS is an aneuploidy caused by trisomy of chromosome 21 and is the leading genetic cause of intellectual disability (Roper & Reeves, 2006). DS is associated with craniofacial dysmorphologies that impair vital functions such as breathing, eating, and speaking. DS craniofacial phenotype is described as brachycephalic, with maxillary hypoplasia leading to facial flatness, depressed nasal bridge and reduced airway passages (Starbuck et al., 2013); dysplastic ears with lobe absence; and eyes with oblique palpebral fissures, epicanthal folds, strabismus and nystagmus (Jones et al., 2021; Korayem & Bakhadher, 2019). Oral alterations include open mouth, cleft lip, lingual furrows and protrusion, macroglossia, micrognathia and narrow palate (Hennequin et al., 1999; Oliveira et al., 2008).

Within RD, we included Morquio syndrome type A (MS, OMIM 253000) because Colombia presents one of the highest prevalence of MS in the world, probably as a result of founder effects. The worldwide prevalence, including other Latin-American countries, ranges from 1 case per 75,000 to 200,000 live births, whereas in Colombia the prevalence rises up to 0.68 per 100,000 live births (Pachajoa et al., 2021). Morquio syndrome is a subtype of Mucopolysaccharidosis disorder caused by more than 180 autosomal recessive mutations in the *GALNS* gene (Herrera et al., 2017) that alter the metabolism of the extracellular matrix glycosaminoglycans (Sawamoto et al., 2020). Keratan and chondroitin sulfate alterations cause irreparable damage to leukocytes and fibroblasts and the typical alterations of MS thus involve the supporting tissue and the osteoarticular system (Suárez-Guerrero et al., 2016). Individuals with MS display skeletal abnormalities such as skeletal dysplasia, short stature and trunk, kyphoscoliosis, pectus carinatum, genu valgum, and joint hyperlaxity (Ortiz-Quiroga et al., 2018). Oral diseases often include periodontal disease, malocclusions, caries and premature tooth loss (Herrera et al., 2017). Individuals with MS show coarse facies with an excessively rapid growth of the head (Suárez-Guerrero et al., 2013). Craniofacial features include a prominent forehead, hypertelorism, prognathism, wide mouth and nose, depressed nasal bridge, plump cheeks and lips with an oversize tongue (Suárez-Guerrero et al., 2013). Dysmorphologies associated with MS vary among individuals, with typical severe phenotypes, although less severe forms have been described as mild or attenuated phenotypes (Suárez-Guerrero et al., 2016).

Finally, we also included in the analyses two RASopathies, Noonan syndrome and Neurofibromatosis type 1, which are prevalent in Valle del Cauca and present altered craniofacial development by genetic mutations that cause Ras/MAPK pathway dysregulation. Noonan syndrome (NS, OMIM 163950) is the most common type of RASopathy and is a rare genetically heterogeneous autosomal dominant disorder caused by mutations in either the *PTPN11, SOS 1, KRAS, BRAF* or *RAF1* genes. The worldwide prevalence of NS is 1 per 1,000 to 1 per 2,500 live births (Hernández-Martín & Torrelo, 2011). Individuals with NS display facial features such as hypertelorism, epicanthic folds, strabismus, downward slanting palpebral fissures, ptosis, high arched palate, deeply grooved philtrum with high peaks of upper lip vermillion border, midfacial hypoplasia and micrognathia, broad flat nose, low-set posteriorly rotated ears, curly/sparse/coarse hair and short webbed neck (Athota et al., 2020).

Neurofibromatosis type 1 (NF1, OMIM 162200) is an autosomal dominantly inherited neurocutaneous disorder caused by a mutation in the *neurofibromin* gene. NF1 has a worldwide incidence of 1 in 2,500 to 1 in 3,000 individuals. The clinical manifestations in NF1 are variable, even within the same family group, and the timing of the onset has a major influence (Hernández-Martín & Torrelo, 2011). Regarding craniofacial traits, individuals with NF1 present macrocephaly, facial asymmetry caused by dysplasia of the sphenoid wings (Khosrotehrani et al., 2003), as well as bone deformities caused by plexiform neurofibromas, enlarged mandibular canal, retrognathic mandible and maxilla, and short cranial base (Visnapuu et al., 2018).

To assess the facial phenotypes associated with these disorders, individuals diagnosed with DS, MS, NS and NF1 and age matched controls were recruited for photographic sessions at educational and research centers in Cali (Colombia) in 2021. The photographic material was taken under the protocol approved by ethics committee “Human Research Ethics Committee of the Icesi University” with Approval Act No. 309”. To photograph the participants and to record relevant clinical information, we obtained informed consent from the participants or from their parents or legal guardians in the case of minor children, in accordance with national guidelines and regulations.

### Facial image acquisition and anatomical landmark collection

Facial shape was captured from 2D images taken using a professional digital camera (SONY Alpha 58 +18-55) that was attached to a tripod and placed at one-meter distance in front of the participants. To capture a natural facial gesture, the images were acquired in an upright position with facial neutral expression.

To measure facial shape of each individual and to detect the traits associated with each disorder, we recorded the 2D coordinates of a set of 18 anatomical facial landmarks (Figure 1 and Supplementary Table 1). Landmarks were acquired using an automatic facial landmark detection procedure adapted from the open-source software library Dlib (King, 2009). From the set of landmarks registered by Dlib, 15 landmarks directly matched our configuration of 18 facial landmarks (Figure 1). Three additional landmarks were approximated through direct computations between the landmarks coordinates automatically returned by Dlib: the glabella was computed as the midpoint point between the innermost points located in the eyebrows, and the palpebrale inferius landmarks of the right and left eyes were computed as the midpoint between the two central lower eyelid landmarks. The validity of the data was assessed by comparing the coordinates of landmarks automatically detected by Dlib to the coordinates of landmarks manually collected by an expert facial morphologist in a set of 20 facial images. The average deviation measured as the root mean square error (RMSE) was 2 mm, which falls within acceptable measurement error standards (Stull et al., 2014).

**Figure 1.**
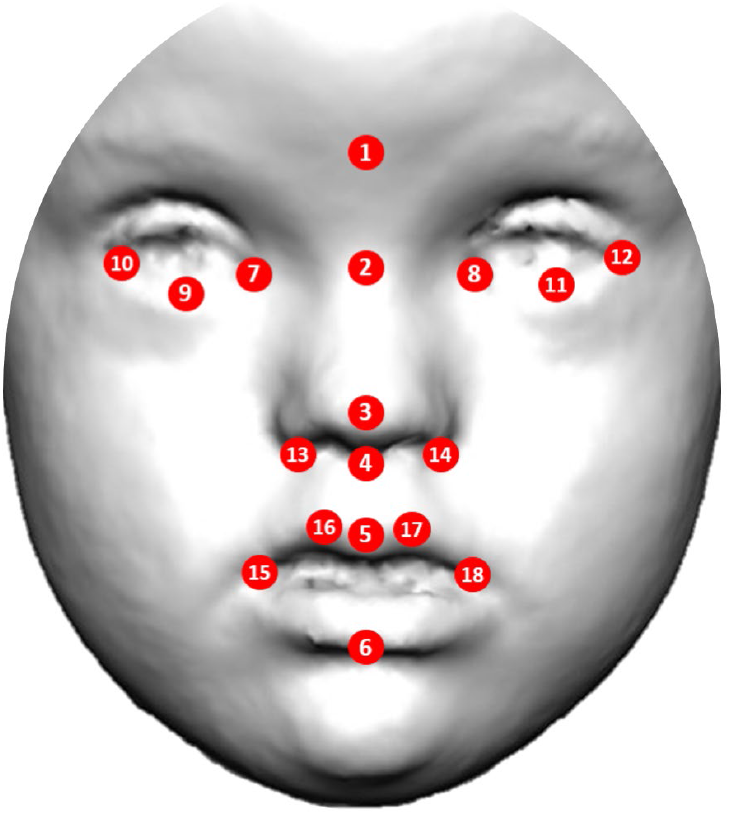
Anatomical position of facial landmarks used in morphometric and statistical analyses to quantify dysmorphologies associated to genetic and rare disorders Down, Morquio, Noonan syndromes and Neurofibromatosis type 1 in a Colombian population.

After inspection of landmark coordinates for gross error, anatomical landmark coordinates were used to conduct morphometric analyses.

### Quantification of facial phenotypes

We used Euclidean distance matrix analysis (EDMA) to describe the facial phenotype associated to each syndrome and to detect specific facial traits that were significantly different between controls and individuals diagnosed with DS, MS, NS and NF. EDMA is a robust morphometric method for assessing local differences between samples (Lele & Richtsmeier, 1991). EDMA detects which linear distances significantly differ between pairwise sample contrasts and compares patterns of significant differences across samples.

First, to account for size differences between subjects, the 2D coordinates of the facial landmarks of each subject were scaled by their centroid size, estimated as the square root of the sum of squared distances of all the landmarks from their centroid (Rohlf & Slice, 1990). After scaling, as EDMA represents shape as a matrix of linear distances between all possible pairs of landmarks, a total of 153 unique facial measurements were calculated for each individual. Linear distances were compared for each group of DS, MS, NS and NF syndromes with control individuals by performing a two-tailed two-sample shape contrasts on all unique inter-landmark linear distances from each sample. Statistical significance was assessed using a non-parametric bootstrap test with 10,000 resamples. EDMA statistically evaluated the number of significant local linear distances in each two-sample comparison based on confidence interval testing (α = 0.10). Relative differences between patients and controls were computed as (mean distance in controls -mean distance in patients) / mean distance in controls. To pinpoint specific local shape differences and to reveal the unique morphological pattern of variation associated with each disorder, the ten longest and shortest significant relative differences were plotted on facial figures.

### Facial dysmorphology score

To confirm that the results were not random due to the small sample sizes available in rare diseases, we combined the results from EDMA with an iterative bootstrapping method that further assessed whether the facial dysmorphologies associated to each syndrome were statistically significant (Starbuck et al, 2021). First, we estimated from the EDMA results a facial dysmorphology score (FDS) as the percentage of significantly different distances between patient and control groups. Then, we ran simulations with random samples of controls and patients generated by iterative bootstrapping to assess the statistical significance of the patterns revealed by EDMA. For each disorder, we first created subsamples of N randomly chosen controls (where N is the total number of patients available in the sample). Then, using a subsampling approach, we automatically generated random pseudo-subsamples containing a known number of patients (namely M). This procedure was repeated with increasing numbers of patients and resulted in a series of staggered pseudo sub-samples that contained from M = 0 to M = N patients. In each of these simulations, we computed an EDMA analysis and an FDS score. The results from each round of random groups were separately represented in histograms. Finally, we compared the distribution of FDS random values with the FDS observed in the whole sample. The *P*-value assessing the statistical significance of the comparison (α = 0.05) was computed as the ratio between the number of simulations containing no patients that provided a higher FDS than the observed FDS and the total number of simulations.

### Face2Gene diagnostic assessment

To assess the accuracy of automated diagnostic methods in the Colombian sample, we compared the clinical diagnosis based on genetic testing with the diagnosis estimated by the Face2Gene technology (FDNA Inc., Boston, MA, USA) using the frontal facial 2D images. Following Gurovich et al. (2019), we assessed the top-one and top-five accuracies for each disorder, estimated as the percentage of cases where the Face2Gene model predicted the correct syndrome as the first result or within the five first results from the sorted list of probable diagnosis. We also calculated these accuracies expanding the diagnostic range to the disorder family.

Moreover, we evaluated the similarity between the facial Gestalt models used by Face2Gene for syndrome classification and the Colombian individuals with clinical diagnosis. For each individual, we selected the first diagnostic prediction that matched their clinical and genetic diagnosis and recorded the Gestalt similarity. We classified the level of similarity between the individual and the corresponding Gestalt model into seven categories, including “very low”, “low”, “low-medium”, “medium”, “medium-high”, “high”, and “very high” Gestalt similarity, using the “gestalt level” barplot provided by Face2Gene.

## RESULTS

EDMA analyses showed that each syndrome presented a characteristic facial phenotype. In individuals with Down syndrome, all facial structures including the eyes, nose and mouth presented significant differences as compared to controls. Overall, DS was associated with wider but shorter facial traits (Figure 2A).

**Figure 2.**
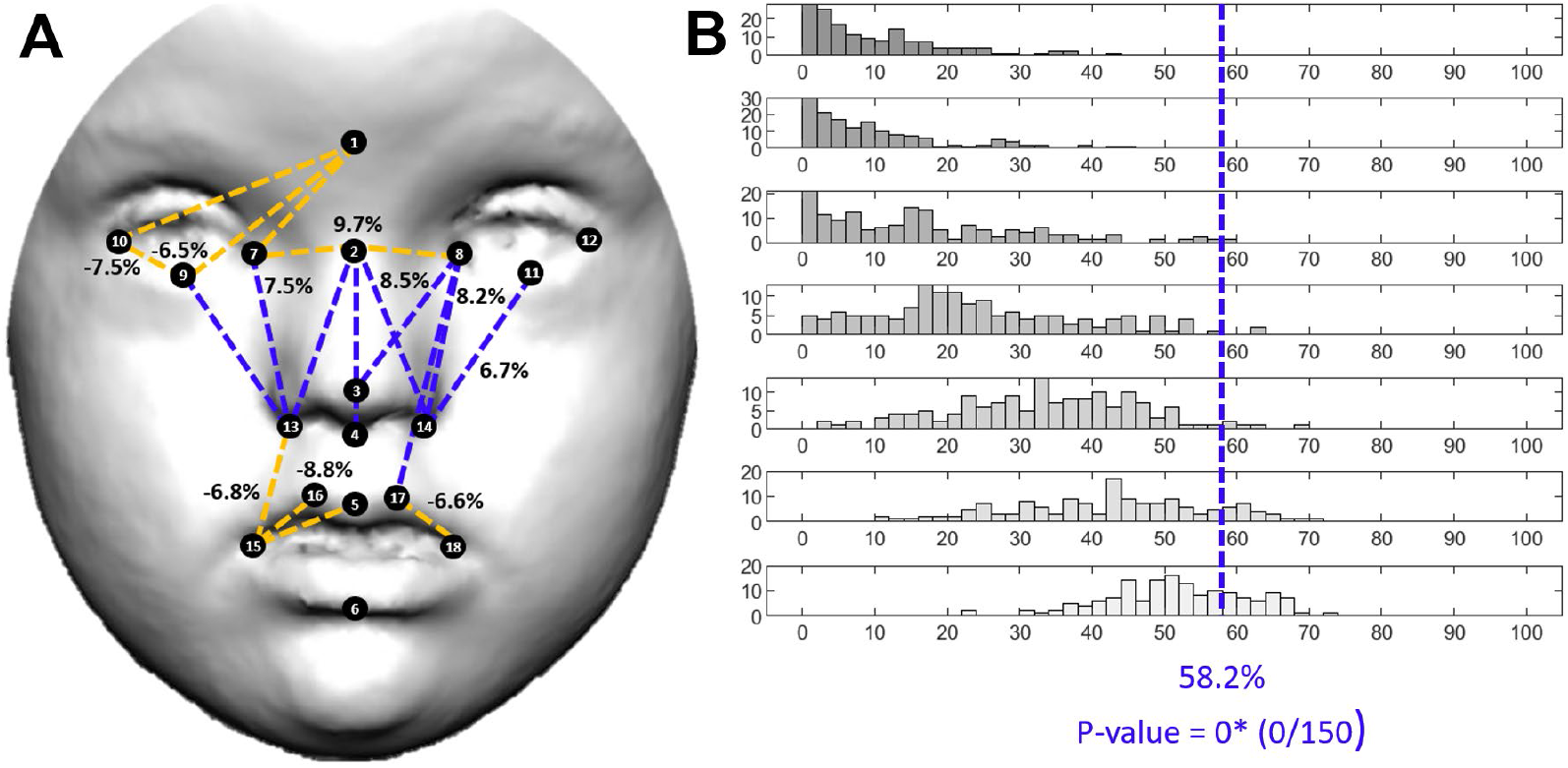
Localized Euclidean Distance Matrix Analysis facial shape pairwise contrasts and iterative bootstrapping tests of facial dysmorphology between controls and individuals diagnosed with Down syndrome. A) EDMA results. Dotted lines represent facial measurements significantly different in control and patient groups. Blue lines indicate measurements that are shorter in patients as compared to controls, whereas orange lines represent measurements that are longer in patients. B) Iterative bootstrapping tests based on facial dysmorphology scores (FDS). Histograms represent the simulation results for each random group separately, which contain an increasing number of patients, from no patients (M = 0) to all patients (M = N). The blue line shows the FDS score obtained with the complete sample of control and patients. * Statistically significant *P*-value.

Results showed 6.5% increase of relative distance between the midpoint between the eyebrows (glabella) and the most inferior medial point of the lower right eyelid (palpelabre inferius), and 7.5% increase between the right palpelabre inferius and the outer commissure of the right eyes (exocanthion), indicating hypertelorism. Additionally, in this Colombian sample, people with DS exhibited longer measurements in the buccal portion, with a 6-8% increase of mouth width as measured from the crista philtri to the chelions (Figure 2A). However, the midfacial and nasal regions were reduced (Figure 2A). People with DS presented a 6-8% reduction in measurements of midfacial height, with the largest difference detected as a 9.7% reduction of the distance between the tip and the root of the nose (Figure 2A). The facial dysmorphology score (FDS) indicated that up to 58.2% of facial traits were significantly different in people with DS (Figure 2B).

The facial pattern associated with Morquio syndrome was also characterized by wider and shorter midfacial traits, as observed in Down syndrome. However, facial dysmorphologies were more abundant and severe in MS than in DS, with 65.4% of facial traits significantly different in diagnosed individuals and higher percentages of relative change (Figure 3 A, B).

**Figure 3.**
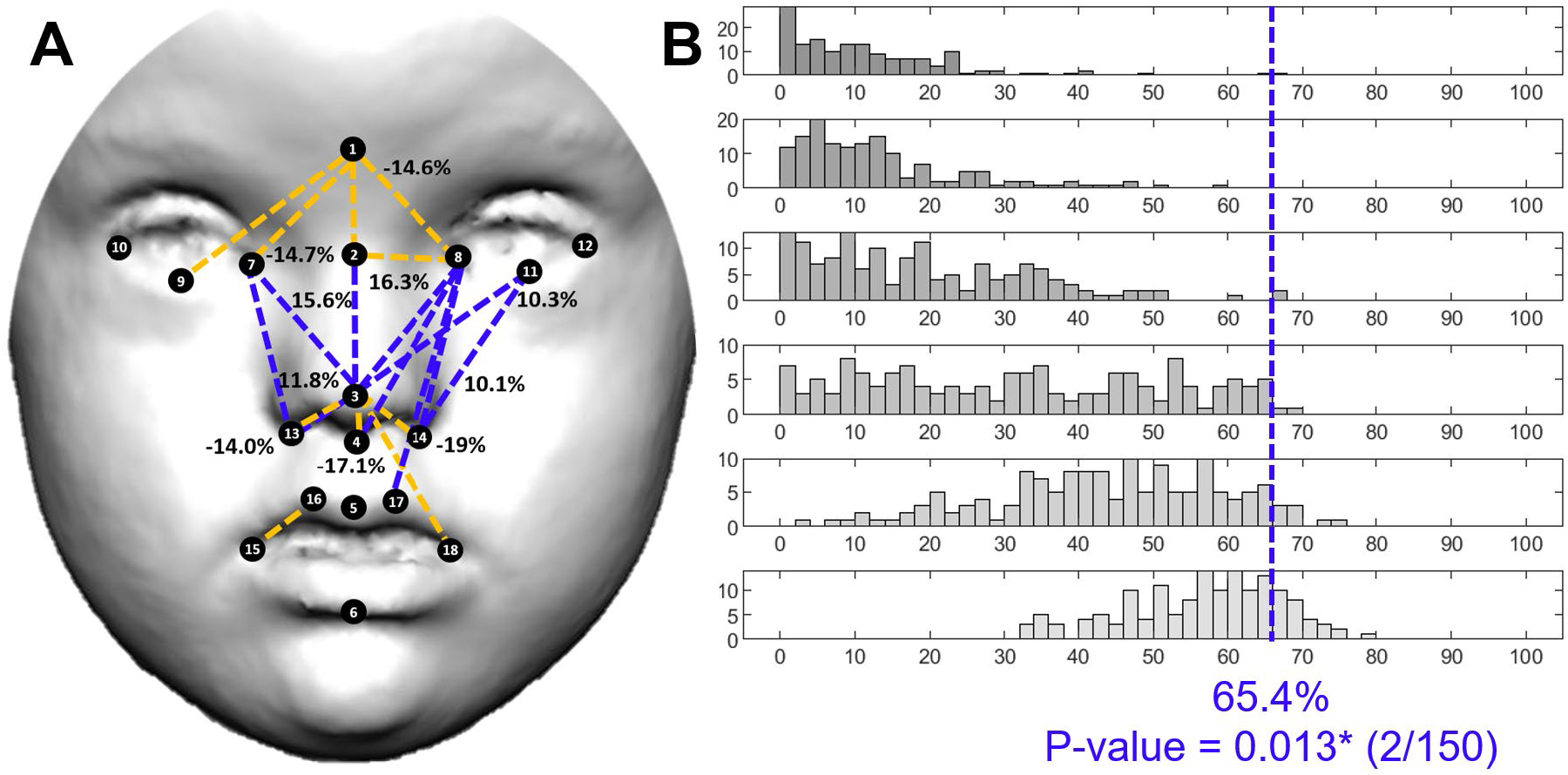
Localized Euclidean Distance Matrix Analysis facial shape pairwise contrasts and iterative bootstrapping tests of facial dysmorphology between controls and individuals diagnosed with Morquio syndrome. For details see legend in Figure 2.

The most affected regions were the midface and the nose, whereas the mouth was the least affected. Individuals with MS presented hypertelorism, with 14% increase in the distance between the midpoint between the eyebrows (glabella) and the inner commissures of the left and right eyes (endocanthions), even larger compared to the differences observed in DS. Individuals with MS also showed larger distances in the base of the nose, with 14-19% increase in the distance from the tip of the nose to the insertion of the right and left alar bases (subalare) as compared to controls. Mouth width was also increased in MS. On the contrary, midfacial heights measuring the distance between the eyes and the nose were significantly reduced from 10 to 16% in individuals with MS (Figure 3A).

In Noonan syndrome, facial dysmorphologies were abundant and concentrated in the orbital and nasal regions. EDMA detected significantly increased distances in the upper face, but decreased distances in the midface (Figure 4A). Patients presented a lower position of the eyes, with 9 to 13% increased distances between the glabella or sellion and the landmarks located in the eyes. The mouth also showed a more inferior position, with 8-10% increased relative distance between the tip of the nose and the superior lip, but the shape of the mouth did not show large differences between patients and controls. The reduction of midfacial heights in individuals with NS ranged from 5 to 11%, with a similar magnitude as in DS (Figure 4A). FDS indicated that 47.7% of facial traits were significantly different in NS (Figure 4B).

**Figure 4.**
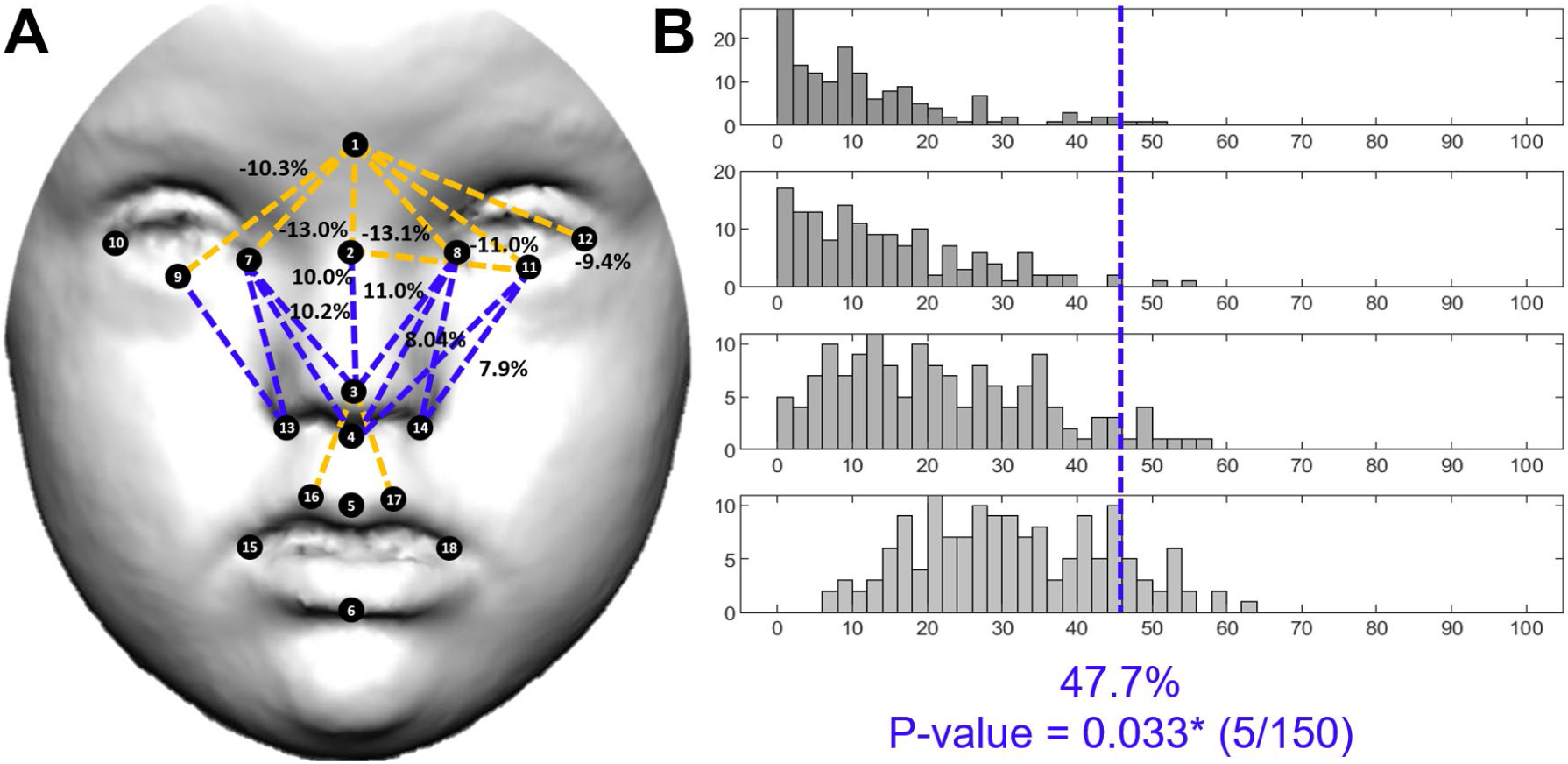
Localized Euclidean Distance Matrix Analysis facial shape pairwise contrasts and iterative bootstrapping tests of facial dysmorphology between controls and individuals diagnosed with Noonan syndrome. For details see legend in Figure 2.

Neurofibromatosis type 1 was associated with minor facial dysmorphologies, which were less abundant and less severe than the previous syndromes (Figure 5A). Individuals with NS only presented 11.4% of significantly different facial traits as compared to controls and the percentages of relative change were low, mostly ranging from 1 to 5% (Figure 5A, B). The largest difference was a 10% increase in facial distance between the glabella and the labiale superius (Figure 5A). Along with larger distances in the midline of the face, EDMA detected reduced distances on the right and left sides of the face, with shorter distances from the right and left chelion to the eye landmarks, the endocanthion and the palpebrale inferius. Hypertelorism was not present in individuals with NF1 (Figure 5A). In NF1, the FDS score was not significant (Figure 5B), indicating that the facial dysmorphology pattern associated with NF1 is so subtle that overall is not larger than facial differences that can be randomly detected within control individuals.

**Figure 5.**
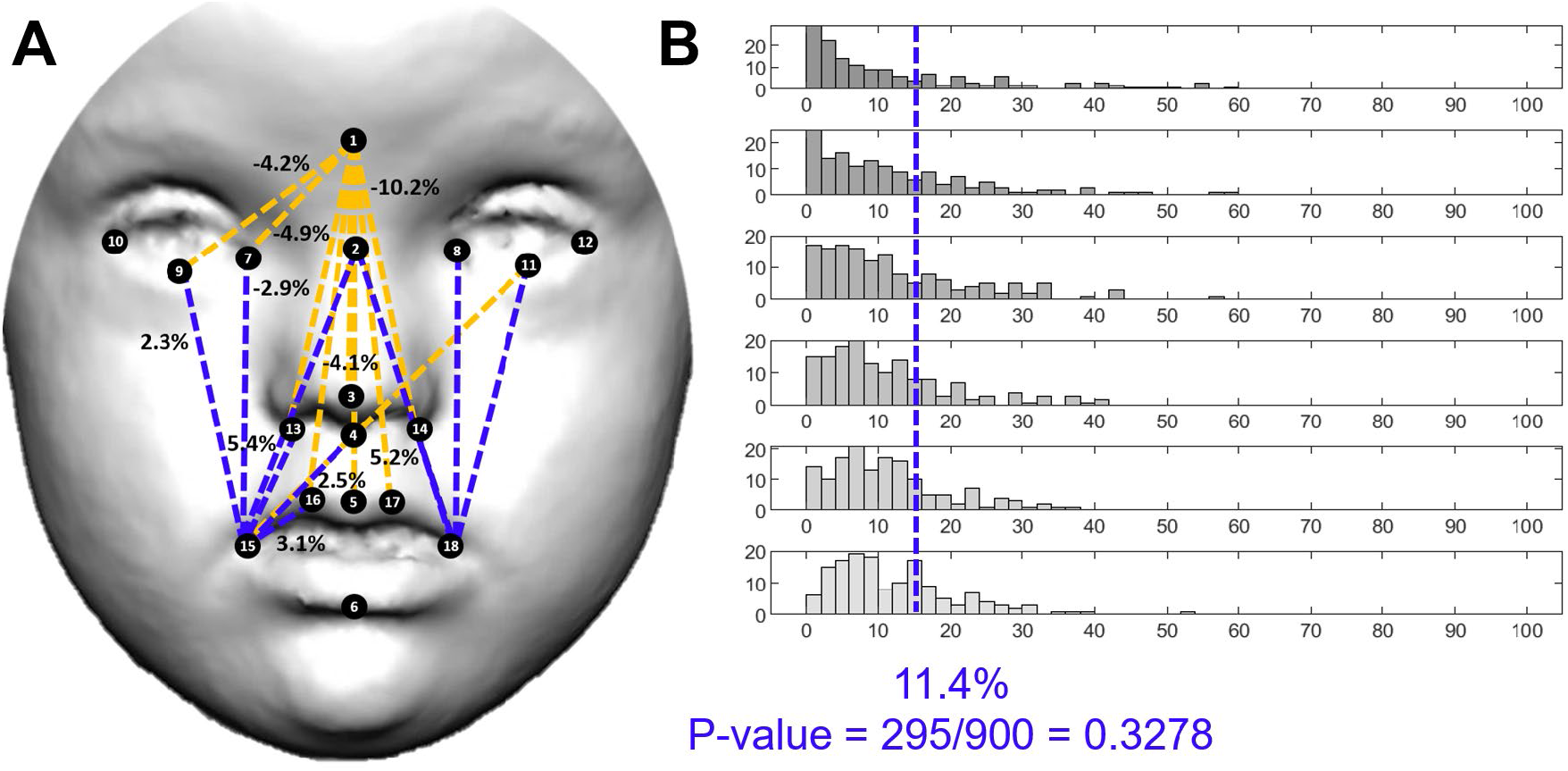
Localized Euclidean Distance Matrix Analysis facial shape pairwise contrasts and iterative bootstrapping tests of facial dysmorphology between controls and individuals diagnosed with Neurofibromatosis type 1. For details see legend Figure 2.

Nevertheless, the simulation tests demonstrated that the facial dysmorphologies measured as FDS scores on individuals diagnosed with Down, Morquio and Noonan syndromes were significant and different from random comparisons in control individuals. Few simulations resulted in a higher FDS than the FDS obtained with the complete sample (Figures 2B, 3B, 4B, first row and blue line). Moreover, in DS, MS and NS, facial dysmorphology scores increased as larger numbers of diagnosed individuals were included in the simulations (Figures 2B, 3B, 4B, middle rows), confirming the severity of the facial dysmorphologies associated to these syndromes. Finally, the simulations comparing all recruited diagnosed individuals with random subsamples of control individuals (Figure 2B, 3B, 4B, last row) indicated that FDS scores can range widely from 10 to 80%, underscoring the biasing effects of control samples.

After quantifying the facial dysmorphologies associated to DS, MS, NS and NF1 in the Colombian sample, we tested the accuracy of the diagnosis provided by the automatic diagnostic algorithms of Face2Gene. We assessed the correspondence between the estimated Face2Gene diagnosis based on facial frontal 2D images with the diagnosis made by clinicians based on genetic testing.

Face2Gene estimated Down syndrome diagnosis with top-1 accuracy of 100%, as DS diagnosis was listed as the first diagnosis in all individuals with an average Gestalt similarity of 6.2 (Table 2). In 79% of individuals, DS diagnosis was associated with very high Gestalt similarity values, but in 21% of individuals the Gestalt similarity was lower, ranging from medium-high to very low values (Table 2, Figure 6).

**Table 2.** Accuracy of Face2Gene diagnosis based on 2D facial images in Down, Morquio, Noonan and Neurofibromatosis type 1 syndromes in a Colombian population. % Of cases matching the genetic diagnosis are provided for each syndrome, as well as Gestalt similarity values.

|  | Top-1 accuracy |  |  |  | Top-5 accuracy |  |  |  |
| --- | --- | --- | --- | --- | --- | --- | --- | --- |
|  | Exact diagnosis |  | Within disorder family |  | Exact diagnosis |  | Within disorder family |  |
|  | % cases | Gestalt similarity | % cases | Gestalt similarity | % cases | Gestalt similarity | % cases | Gestalt similarity |
| **DS** | 100 | 6.2 | 100 | 6.2 | 100 | 6.2 | 100 | 6.2 |
| **MS** | 0 | 0 | 36.4 | 5.6 | 45.4 | 3 | 100 | 3.4 |
| **NS** | 66.7 | 5.2 | 66.7 | 5.2 | 77.8 | 4.7 | 88.9 | 4.4 |
| **NF1** | 8.3 | 1 | 50 | 1.7 | 66.6 | 1.2 | 66.6 | 1.6 |

**Figure 6.**
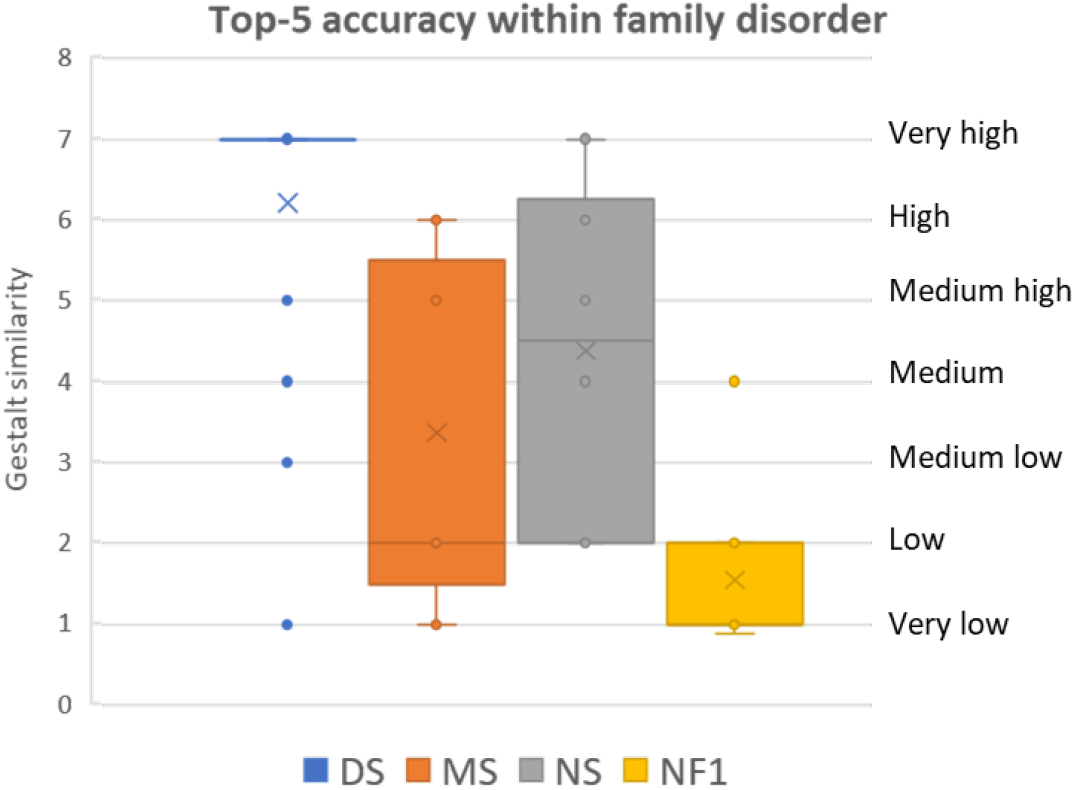
Gestalt similarity scores between diagnosed individuals and Face2Gene gestalts in Down, Morquio, Noonan and Neurofibromatosis type 1 syndromes. Histograms based on top-5 accuracy Face2Gene predictions within family disorder.

On the contrary, the top-1 accuracy of Morquio syndrome was 0%, as Face2Gene never listed the specific diagnostic of mucopolysaccharidosis type IVA (MPSIVA) as a first prediction (Table 2). Although Face2Gene could not identify the specific type of MS, the automatic diagnostic algorithms associated the facial dysmorphologies with a diagnosis related with mucopolysaccharidosis disorders in 36.4% of cases, with a medium-high average Gestalt similarity of 5.6 (Table 2). When the first 5 diagnostic predictions were considered, the top-5 accuracy raised to 45.4% for exact MPSIVA diagnosis and to 100% for mucopolysaccharidosis disorders, but with a low-medium Gestalt similarity (Table 2, Figure 6).

The top-1 accuracy of Face2Gene for Noonan syndrome was 66.7%, with a medium-high average Gestalt similarity of 5.2 (Table 2). Top-5 accuracy increased to 77.8% for exact NS diagnosis, and to 88.9% when considering Noonan Syndrome-Like Disorder diagnoses, with a medium Gestalt similarity of 4.4 and wide variation among individuals (Table 2, Figure 6).

Finally, in Neurofibromatosis type 1, Face2Gene presented a top-1 accuracy of 8.3% associated with a very low Gestalt similarity of 1 (Table 2). When diagnoses within the RASopathies disorder family were considered, the top-1 accuracy raised to 50%, as 5 out of 12 individuals were diagnosed as Noonan syndrome (Table 2). The top-5 accuracy was 66.6% and was associated with low Gestalt similarity values of between 1 and 2 in 87.5% of individuals (Table 2, Figure 6).

## DISCUSSION

Our analyses provided an accurate quantitative comparison of facial dysmorphologies in Down, Morquio and Noonan syndromes, as well as in Neurofibromatosis type 1, in a Latin-American population from Colombia. An objective and highly detailed description of the facial phenotype is a major improvement over qualitative descriptions of the complex facial dysmorphologies associated with these genetic disorders. We quantified local facial trait differences presented in people diagnosed with these disorders as compared to age matched controls of the same population, localizing the largest statistically significant facial dysmorphologies.

Our results indicated differential facial patterns associated with each disorder, with major significant dysmorphologies in Down, Morquio and Noonan syndromes, and minor facial dysmorphologies associated with NF1. Different types of genetic alterations, which ranged from aneuploidy and overall genetic imbalance as in DS; to point genetic mutations affecting different processes, such as the metabolism of mucopolysaccharides in MS; or signaling pathways, such as the *RAS/MAPK* pathway in NS and NF1, significantly affected the facial phenotypes. In genetic and rare diseases, genetic alterations deviate the signaling pathways regulating normal facial development (Jones et al., 2021; Terrazas et al., 2017), altering normal morphogenesis and growth during pre- and postnatal development (Kouskoura et al., 2011).

### Population-specific facial traits in Colombian individuals with rare disorders

Overall, the facial patterns associated to each syndrome in the Colombian Latin-American population coincide with the descriptions reported in the literature (Khosrotehrani et al., 2003; Suárez-Guerrero et al., 2013; Starbuck et al., 2013; Athota et al., 2020). However, there are specific local traits that differ, suggesting that facial traits associated to disease might be modulated by population ancestry and influenced by different evolutionary and adaptive histories of human populations (Kruszka et al., 2017).

In Down syndrome, we found a facial dysmorphologies that are consistent with the patterns reported in the literature for populations of European descent. Our analyses detected differences in linear facial measurements that correspond to typical DS traits such as hypertelorism, maxillary hypoplasia and shorter and wider faces associated to a brachycephalic head (Jones et al., 2021; Korayem & Bakhadher, 2019). Results also suggested other characteristic traits of DS, such as midfacial retrusion and depressed nasal bridge (Starbuck et al., 2013). Open mouth and macroglossia (Hennequin et al., 1999; Oliveira et al., 2008) were also observed during the photographic sessions in the participants of our study. However, in contrast to European and North American populations (Starbuck et al., 2021), in the Colombian population we detected that the mouth was wider in individuals diagnosed with DS as compared to euploid controls. This difference may be caused by unnatural facial gestures of the participants when asked to close the mouth during the photo shoot, or by facial differences associated to ancestry. Indeed, Kruszka et al., (2017) analyzed individuals diagnosed with DS in diverse populations and showed craniofacial differences between individuals from different ethnicities (Africans, Asians, and Latin Americans), demonstrating that ancestry is a relevant factor when assessing craniofacial variation associated to rare diseases.

In Morquio syndrome, the facial dysmorphologies observed in Colombian individuals were consistent with traits reported in the literature, which included hypertelorism, prognathism, wide nose and wide mouth (Hung et al., 2016 Suárez-Guerrero et al., 2013; Herrera et al., 2017). In our pediatric cohort from Colombia, Morquio syndrome was associated with the most severe facial dysmorphologies. Considering that keratan and chondroitin sulfate alterations associated with MS accumulate over life and cause irreparable damage to the osteoarticular system, facial dysmorphologies associated with MS are expected to increase with age due to an excessively rapid growth of the head, becoming even more severe in adult individuals (Suárez-Guerrero et al., 2013). Further research is required to assess this hypothesis and to test whether pharmacological treatments can slow down the progression of the disease and reduce the facial dysmorphologies associated with MS. This is especially relevant in Colombia, a country with one of the highest prevalence of MS in the world (Pachajoa et al., 2021).

In Noonan syndrome, we detected hypertelorism, downward slanting palpebral fissures, and midfacial hypoplasia in the Colombian population, as reported in populations of European descent (Athota et al., 2020). Moreover, our results quantified changes in the position of the mouth in Colombian individuals diagnosed with NS not reported before (Lores et al., 2020).

The facial pattern associated with NF1 in individuals from Colombia was also compatible with some typical traits of NF1, such as midface hypoplasia (Hernández-Martín & Torrelo, 2011). However, our results did not underscore facial asymmetry or hypertelorism as prominent facial differences between diagnosed individuals and controls in the Colombian population (Hernández-Martín & Torrelo, 2011).

Comparative quantitative studies from different world regions are not usually available for most genetic and rare disorders, and reference data for diagnosis is mainly based on phenotypes defined on populations of European descent. In fact, almost no images of individuals of Latin American origin are included in reference medical texts such as Smith’s Recognizable Patterns of Human Malformation (Jones et al., 2021). However, previous studies have reported that in Noonan and other genetic syndromes, such as Turner, 22q11.2 and Cornelia de Lange syndromes presented distinctive facial traits that were population specific, with clinical features that were significantly different in Africans, Asians, and Latin Americans (Kruszka et al., 2017). Our results in a Colombian population further support this evidence, highlighting the need to extend the analyses to populations from all over the world to achieve a complete and more accurate phenotypic representation of genetic and RD to optimize the diagnostic potential of facial biomarkers in the clinical practice.

### Reduced accuracy diagnosis in a Colombian population with diverse ancestry

Deep learning algorithms such as Face2gene have shown potential as a reliable and precise tool for genetic diagnosis by image recognition (Gurovich et al., 2019; Hsieh et al., 2021; Park et al., 2022; Pascolini et al., 2022). In the Colombian cohort analyzed here, Face2gene diagnosed with 100% accuracy Down syndrome, which is one of the most common and more accurately represented genetic disorder. In Noonan syndrome, the percentage of top1-accuracy was lower (66.7%), but still correctly identified the disorder in the majority of individuals when considering the top5-accuracy within Noonan syndrome-like disorders (88.9%). However, in Morquio syndrome, despite being associated with the most severe facial dysmorphologies, the top1-accuracy for exact diagnosis of Mucopolysaccharidosis type IVA was 0% and a low percentage of cases (36.4%) were diagnosed with a Mucopolysaccharidosis-like syndrome as the first prediction.

Interestingly, there was a wide range of variation in gestalt similarity for most disorders, even for Down syndrome. Visual inspection of cases suggested that most individuals associated with the lowest gestalt similarity scores were highly admixed, with diverse contributions of Amerindian, African and European ancestry components. Future analyses need to investigate the role of population ancestry and further assess the reliability and validity of automatic diagnostic tools in admixed populations from non-European descent. As evidenced in this study, where syndromes such as MS, NS, NF1 were associated with low accuracy scores versus more frequent syndromes such as DS, this is critical in rare syndromes with heterogenous clinical presentation and phenotype, where clinical diagnosis is a challenging process (Schieppati et al., 2008; Bannister et al., 2020) that may take several years, leading to the so-called diagnostic odyssey (González-Lamuño & García Fuentes, 2008).

Precise and early diagnosis of genetic and rare disorders is crucial for adequate health care and clinical management. Without a diagnosis, individuals and their families must proceed without basic information regarding their health and future developmental outcomes (Bannister et al., 2020). Even though gene-based technologies have greatly improved diagnostic procedures (Agbolade et al., 2020), the mutations causing many rare diseases are still unknown and access to genetic testing is limited (Suárez-Obando, 2018). Genetic consultations may become a long process, and broad molecular testing such as exome and genome sequencing represent a high expense that is not affordable for all families and health care systems, especially in low-medium income countries (González-Lamuño & García Fuentes, 2008).

In this context, faster, non-invasive and low-cost diagnostic methods based on facial phenotypes emerge as complementary tools for providing earlier first reliable diagnoses (Gurovich et al., 2019; Agbolade et al., 2020; Hallgrímsson et al., 2020; Hsieh et al., 2021), especially in low-middle income countries with low budget and resources for molecular testing. Qualitative visual assessment of craniofacial dysmorphology is frequently employed for diagnosis, clinical management and treatment monitoring (Jones et al., 2021). Experts in dysmorphologies can identify the facial “gestalt” distinctive of many dysmorphic syndromes (Jones et al., 2021). However, this facial assessment relies on the expertise of the clinician and is very challenging, as there is no clear one-to-one correspondence between disorders and facial dysmorphologies. Different genetic mutations can cause the same syndrome or similar phenotypes, whereas the same mutation can induce different phenotypes (Aldridge et al., 2010; Martínez-Abadías et al., 2011). In addition, within the same rare disease there may be several subtypes, and symptoms may vary even within individuals of the same genetic disorder and the same family (Suárez-Obando, 2018). This complex biology generates confusion at the time of diagnosis and warrants the development of efficient, objective and reliable diagnostic methods.

Computer-assisted phenotyping can overcome these pitfalls and provide widely accessible technologies for quick syndrome screening (Bannister et al., 2020). In this automated approach, methods can be based on 2D or 3D images (Gurovich et al., 2019; Hallgrímsson et al., 2020; Hsieh et al., 2021). The advantage of 2D methods is that data collection is easy and can be readily translated into the clinical practice, as physicians can take facial images even with simple digital cameras or smartphones. The collection of 3D models is more sophisticated and requires specialized equipment but provides more accurate phenotype descriptions by incorporating the depth dimension.

To further improve the methods of craniofacial assessment to diagnose individuals with genetic syndromes and RD that exhibit facial dysmorphologies, it is crucial to assess the large morphological variation displayed by human populations in facial phenotypes. Factors such as age, sex and ancestry should be accounted for in diagnostic methods. Clinical manifestations in some genetic disorders usually begin at an early age, with two thirds of patients expressing symptoms before the second year of birth (Suárez-Obando, 2018), but in other disorders facial dysmorphologies develop later, during postnatal development. Male and female faces present sexual dimorphism at adulthood (Enlow & Hans, 1996) and diseases can differently affect the facial phenotype depending on sex differences (Martínez-Abadías et al., 2021). As suggested here, differences in population ancestry can also significantly influence the facial phenotype and the potential for clinical diagnosis.

Therefore, recruitment of participants must be expanded to include as many individuals with RD as possible, together with large comparative samples of age-matched controls, from both sexes and from all the populations in the world representing varied ancestries. For instance, the population in Southwestern Colombia is characterized by high levels of admixture from people with Native American, African, and European ancestry (Urrea-Giraldo & Álvarez, 2017; Adhikari et al., 2017). Including the morphological variation of faces from such different ancestry backgrounds is key to pinpoint the facial dysmorphologies associated with diseases in worldwide diverse populations (Conley et al., 2017). Our simulation analyses further highlighted the importance of maximizing the recruitment of diagnosed and control individuals, as results considerably changed depending on the cohort and sample sizes

## CONCLUSION

Our results underscore that facial phenotypes associated with genetic and rare disorders are influenced by population ancestry (Kruszka et al., 2017; Dowsett et al., 2019) and that diverse genetic background variation can modulate the phenotypic response to disease, affecting the accuracy of current tools of clinical diagnosis. In the future, deep learning algorithms including a high variety of populations with different ancestry backgrounds will optimize the precision and accuracy of diagnosis in an unbiased approach. Such predictive models will support clinicians in decision-making across the world.

## Data Availability

All data produced in the present study are available upon reasonable request to the authors

## ACKNOWLEDGEMENTS

We are grateful for the voluntary collaboration of all participants, including children and their families. We are thankful to Colegio Ecológico Scout and Universidad Icesi for granting us permission to organize the photographic sessions. We thank Max Rubert for technical photographic assistance. We acknowledge support from Proyecto COL0012168-1097 Interfacultades-ICESI, and 2017SGR1630 Research group (AGAUR).

## LITERATURE CITED

Aase JM. (1990). The physical examination in dysmorphology. In: Diagnostic dysmorphology. New York and London: Plenum Medical Book Company. p 33–42

Adhikari, K., Chacón-Duque, J.C., Mendoza-Revilla, J., Fuentes-Guajardo, M., & Ruiz-Linares, A. (2017). The Genetic Diversity of the Americas. Annual Review of Genomics and Human Genetics, 18:277–296. https://doi.org/10.1146/annurev-genom-083115-022331

Aivazidis, S., Coughlan, C. M., Rauniyar, A. K., Jiang, H., Liggett, L. A., Maclean, K. N., & Roede, J. R. (2017). The burden of trisomy 21 disrupts the proteostasis network in Down syndrome. PloS One, 12(4), e0176307. https://doi.org/10.1371/journal.pone.0176307

Agbolade, O., Nazri, A., Yaakob, R., Ghani, A. A., & Cheah, Y. K. (2020). Down Syndrome Face Recognition: A Review. Symmetry, 12(7), 1182. https://doi.org/10.3390/sym12071182

Aldridge, K., Hill, C. A., Austin, J. R., Percival, C., Martinez-Abadias, N., Neuberger, T., Wang, Y., Jabs, E. W., & Richtsmeier, J. T. (2010). Brain phenotypes in two FGFR2 mouse models for Apert syndrome. Developmental Dynamics, 239(3), 987–997. https://doi.org/10.1002/dvdy.22218

Ardelean, C. F., Becerra-Valdivia, L., Pedersen, M. W., Schwenninger, J.-L., Oviatt, C. G., Macías-Quintero, J. I., Arroyo-Cabrales, J., Sikora, M., Ocampo-Díaz, Y. Z. E., Rubio-Cisneros, I. I., Watling, J. G., de Medeiros, V. B., De Oliveira, P. E., Barba-Pingarón, L., Ortiz-Butrón, A., Blancas-Vázquez, J., Rivera-González, I., Solís-Rosales, C., Rodríguez-Ceja, M., … Willerslev, E. (2020). Evidence of human occupation in Mexico around the Last Glacial Maximum. Nature, 584(7819), 87–92. https://doi.org/10.1038/s41586-020-2509-0

Athota, J. P., Bhat, M., Nampoothiri, S., Gowrishankar, K., Narayanachar, S. G., Puttamallesh, V., Farooque, M. O., & Shetty, S. (2020). Molecular and clinical studies in 107 Noonan syndrome affected individuals with PTPN11 mutations. BMC Medical Genetics, 21(1), 50. https://doi.org/10.1186/s12881-020-0986-5

Bannister, J. J., Crites, S. R., Aponte, J. D., Katz, D. C., Wilms, M., Klein, O. D., Bernier, F. P. J., Spritz, R. A., Hallgrímsson, B., & Forkert, N. D. (2020). Fully Automatic Landmarking of Syndromic 3D Facial Surface Scans Using 2D Images. Sensors, 20(11), 3171. https://doi.org/10.3390/s20113171

Becerra-Valdivia, L., & Higham, T. (2020). The timing and effect of the earliest human arrivals in North America. Nature, 584(7819), 93–97. https://doi.org/10.1038/s41586-020-2491-6

Carlson, B.M. (2019). Human embryology & developmental biology. St Louis (Missouri), United States: Elsevier.

Cortés, Fanny. (2015). Las enfermedades raras. Revista Médica Clínica Las Condes. 26. 425–431. https://doi.10.1016/j.rmclc.2015.06.020.

Conley, A. B., Rishishwar, L., Norris, E. T., Valderrama-Aguirre, A., Mariño-Ramírez, L., Medina-Rivas, M. A., & Jordan, I. K. (2017). A Comparative Analysis of Genetic Ancestry and Admixture in the Colombian Populations of Chocó and Medellín. G3 (Bethesda, Md.), 7(10), 3435–3447. https://doi.org/10.1534/g3.117.1118

Castro E Silva, M. A., Ferraz, T., Bortolini, M. C., Comas, D., & Hünemeier, T. (2021). Deep genetic affinity between coastal Pacific and Amazonian natives evidenced by Australasian ancestry. Proceedings of the National Academy of Sciences of the United States of America, 118(14), e2025739118. https://doi.org/10.1073/pnas.2025739118

Dowsett, L., Porras, A. R., Kruszka, P., Davis, B., Hu, T., Honey, E., Badoe, E., Thong, M., Leon, E., Girisha, K. M., Shukla, A., Nayak, S. S., Shotelersuk, V., Megarbane, A., Phadke, S., Sirisena, N. D., Dissanayake, V. H. W., Ferreira, C. R., Kisling, M. S., … Krantz, I. D. (2019). Cornelia de Lange syndrome in diverse populations. American Journal of Medical Genetics Part A, 179(2), 150–158. https://doi.org/10.1002/ajmg.a.61033

Enlow, D.H., & Hans, M.G. (1996). Essentials of facial growth. Saunders Editorial.

Farrera, A., Villanueva, M., Vizcaíno, A., Medina-Bravo, P., Balderrábano-Saucedo, N., Rives, M., Cruz, D., Hernández-Carbajal, E., Granados-Riveron, J., & Sánchez-Urbina, R. (2019). Ontogeny of the facial phenotypic variability in Mexican patients with 22q11.2 deletion syndrome. Head & Face Medicine, 15(1), 29. https://doi.org/10.1186/s13005-019-0213-9

Glasson, E. J., Sullivan, S. G., Hussain, R., Petterson, B. A., Montgomery, P. D., & Bittles, A. H. (2002). The changing survival profile of people with Down’s syndrome: implications for genetic counselling. Clinical Genetics, 62(5), 390–393. https://doi.org/10.1034/j.1399-0004.2002.620506.x

González-José, R., González-Martín, A., Hernández, M., Pucciarelli, H. M., Sardi, M., Rosales, A., & Van der Molen, S. (2003). Craniometric evidence for Palaeoamerican survival in Baja California. Nature, 425(6953), 62–65. https://doi.org/10.1038/nature01816

González-Lamuño, D., & García-Fuentes, M. (2008). Enfermedades de base genética. Anales del Sistema Sanitario de Navarra, 31:105–126.

Gülbakan, B., Özgül, R.K., Yüzbaşıoğlu, A., Kohl, M., Deigner, H., & Özgüç, M. (2016) Discovery of biomarkers in rare diseases: innovative approaches by predictive and personalized medicine. EPMA Journal, 7(24), 1–6. https://doi.org/10.1186/s13167-016-0074-2

Gurovich, Y., Hanani, Y., Bar, O., Nadav, G., Fleischer, N., Gelbman, D., Basel-Salmon, L., Krawitz, P. M., Kamphausen, S. B., Zenker, M., Bird, L. M., & Gripp, K. W. (2019). Identifying facial phenotypes of genetic disorders using deep learning. Nature Medicine, 25(1), 60–64. https://doi.org/10.1038/s41591-018-0279-0

Hammond, P., Hutton, T.J., Allanson, J.E., Campbell, L.E., Hennekam, R.C.M., Holden, S., Patton, M.A., Shaw, A., Temple, I.K., Trotter, M., Murphy, K.C., & Winter, R.M. (2004). 3D analysis of facial morphology. American Journal of Medical Genetics, A 126A:339–348. https://doi:10.1002/ajmg.a.20665

Hammond, P. (2007). The use of 3D face shape modelling in dysmorphology. Archive of Disease in Childhood, 92:1120–1126. https://doi:10.1136/adc.2006.103507

Hammond, P., & Suttie, M. (2012). Large-scale objective phenotyping of 3D facial morphology. Human Mutation, 33:817–825. https://doi:10.1002/humu.22054

Hallgrímsson, B., Percival, C.J., Green. R., Young, N.M., Mio, W., & Marcucio, R. (2015). Morphometrics, 3D Imaging, and Craniofacial Development. Current Topics in Developmental Biology, 115:561–597. https://doi:10.1016/bs.ctdb.2015.09.003

Hallgrímsson, B., Aponte, J. D., Katz, D. C., Bannister, J. J., Riccardi, S. L., Mahasuwan, N., McInnes, B. L., Ferrara, T. M., Lipman, D. M., Neves, A. B., Spitzmacher, J. A. J., Larson, J. R., Bellus, G. A., Pham, A. M., Aboujaoude, E., Benke, T. A., Chatfield, K. C., Davis, S. M., Elias, E. R., … Klein, O. D. (2020). Automated syndrome diagnosis by three-dimensional facial imaging. Genetics in Medicine, 22(10), 1682–1693. https://doi.org/10.1038/s41436-020-0845-y

Hennequin, M., Faulks, D., Veyrune, J-L & Bourdiol, P. (1999). Significance of oral health in persons with Down syndrome: a literature review. Developmental Medicine and Child Neurology, 41(4), 275–283. https://doi:10.1111/j.1469-8749.1999.tb00599.

Hernández Ramírez, I., & Manrique Hernández, R. D. (2006). Prevalencia de síndrome de Down en CEHANI-ESE, San Juan de Pasto Colombia. 1998-2003. Nova, 4(5), 50–56. https://doi.org/10.22490/24629448.347

Hernández-Martín, A., & Torrelo, A. (2011). Rasopathies: Developmental Disorders That Predispose to Cancer and Skin Manifestations. Actas Dermo-Sifiliográficas (English Edition), 102(6), 402–416. https://doi.org/10.1016/j.adengl.2011.02.002

Herrera, L. M. C., Martínez, A. V., López, N. M., Téllez, J. M., & Contreras, X. D. M. (2017). Síndrome de Morquio, enfermedad de interés para la odontopediatría. Presentación de un caso. Revista Pediátrica Electrónica, 14 (4): 2–11.

Hurst, A.C.E. (2018). Facial recognition software in clinical dysmorphology. Current Opinion in Pediatrics, 30:701–706. https://doi:10.1097/MOP.0000000000000677

Johannes, M., Clara, V., Hubert, C., & Raoul, H. (2003). Phenotypic abnormalities: Terminology and classification. American Journal of Medical Genetics, 123A (3), 211–230. https://doi:10.1002/ajmg.a.20249

Jones, K.L., Jones, M.C., & Campo, M. (2021). Smith’s Recognizable Patterns of Human Malformation. Elsevier Health Sciences.

King, D.E. (2009). Dlib-ml: A Machine Learning Toolkit. Journal of Machine Learning Research, 10:1755–1758.

Khosrotehrani, K., Bastuji-Garin, S., Zeller, J., Revuz, J., & Wolkenstein, P. (2003). Clinical Risk Factors for Mortality in Patients With Neurofibromatosis 1: A Cohort Study of 378 Patients. Archives of Dermatology, 139(2). https://doi.org/10.1001/archderm.139.2.187

Köhler, S., Carmody, L., Vasilevsky, N., Jacobsen, J.O.B., Danis, D., Gourdine, J-P., Gargano, M., Harris, N.L., Matentzoglu, N., McMurry, J.A., Osumi-Sutherland, D., Cipriani, V., Balhoff, J.P., Conlin, T., Blau, H., Baynam, G., Palmer, R., Gratian, D., Dawkins, H., Segal, M., Jansen, A.C., Muaz, A., Chang, W.H., Bergerson, J,. Laulederkind, S.J.F., Yüksel, Z., Beltran, S., Freeman, A.F., Sergouniotis, P.I., Durkin, D., Storm, A.L., Hanauer, M., Brudno, M., Bello, S.M., Sincan, M., Rageth, K., Wheeler, M.T., Oegema, R., Lourghi, H., Della Rocca, M.G., Thompson, R., Castellanos, F., Priest, J., Cunningham-Rundles, C., Hegde, A., Lovering, R.C., Hajek, C., Olry, A., Notarangelo, L., Similuk, M., Zhang, X.A., Gómez-Andrés, D., Lochmüller, H., Dollfus, H., Rosenzweig, S., Marwaha, S., Rath, A., Sullivan, K,. Smith, C., Milner, J.D., Leroux, D., Boerkoel, C.F., Klion, A., Carter, M.C., Groza, T., Smedley, D., Haendel, M.A., Mungall, C., & Robinson, P.N. (2019). Expansion of the Human Phenotype Ontology (HPO) knowledge base and resources. Nucleic Acids Research, 47:D1018–D1027. https://doi:10.1093/nar/gky1105

Korayem, M. & Bakhadher, W. (2019). Craniofacial manifestations of Down syndrome: A review of literature. Academia Journal of Scientific Research, https://10.15413/ajsr.2019.0502.

Kouskoura, T., Fragou, N., Alexiou, M., John, N., Sommer, L., Graf, D., Katsaros, C., & Mitsiadis, T.A. (2011). The genetic basis of craniofacial and dental abnormalities. Schweizer Monatsschrift fur Zahnmedizin, 121 7-8, 636–46.

Kruszka, P., Porras, A. R., Addissie, Y. A., Moresco, A., Medrano, S., Mok, G. T. K., Leung, G. K. C., Tekendo-Ngongang, C., Uwineza, A., Thong, M.-K., Muthukumarasamy, P., Honey, E., Ekure, E. N., Sokunbi, O. J., Kalu, N., Jones, K. L., Kaplan, J. D., Abdul-Rahman, O. A., Vincent, L. M., … Muenke, M. (2017). Noonan syndrome in diverse populations. American Journal of Medical Genetics Part A, 173(9), 2323–2334. https://doi.org/10.1002/ajmg.a.38362

Laignier, M. R., Lopes-Júnior, L. C., Santana, R. E., Leite, F. M. C., & Brancato, C. L. (2021). Down Syndrome in Brazil: Occurrence and Associated Factors. International Journal of Environmental Research and Public Health, 18(22), 11954. https://doi.org/10.3390/ijerph182211954

Lele, S. R. & Richtsmeier, J. T. (1991). Euclidean Distance Matrix Analysis: a coordinate-free approach for comparing biological shapes using landmark data. American Journal of Physical Anthropology, 86, 415–27.

Lores, J., Prada, CE., Ramírez-Montaño, D., Nastasi-Catanese, JA., & Pachajoa, H. Clinical and molecular analysis of 26 individuals with Noonan syndrome in a reference institution in Colombia. American Journal of Medical Genetics, Part C. 184(10), 1042–1051. https://doi.org/10.1002/ajmg.c.31869

Martínez-Abadías, N., González-José, R., González-Martín, A., Van der Molen, S., Talavera, A., Hernández, P., & Hernández, M. (2006). Phenotypic evolution of human craniofacial morphology after admixture: A geometric morphometrics approach. American Journal of Physical Anthropology, 129(3), 387–398. https://doi.org/10.1002/ajpa.20291

Martínez-Abadías, N., Heuzé, Y., Wang, Y., Jabs, E. W., Aldridge, K., & Richtsmeier, J. T. (2011). FGF/FGFR Signaling Coordinates Skull Development by Modulating Magnitude of Morphological Integration: Evidence from Apert Syndrome Mouse Models. PLoS ONE, 6(10), e26425. https://doi.org/10.1371/journal.pone.0026425

Martínez-Abadías, N., Hostalet, N., Mariscal-Uceda, L., Gonzàlez, R., González, A., Sevillano, X., Canales-Rodríguez, E., Salgado-Pineda, P., Salvador, R., Pomarol-Clotet, E. and Fatjó-Vilas, M. (2021). Facial Biomarkers Detect Gender-Specific Traits for Bipolar Disorder. The FASEB Journal, 35: https://doi.org/10.1096/fasebj.2021.35.S1.03695

Nguengang Wakap, S., Lambert, DM., Olry, A., Rodwell, C., Gueydan, C., Lanneau, V., Murphy, D., Le Cam, Y., & Rath, A. (2020). Estimating cumulative point prevalence of rare diseases: analysis of the Orphanet database. European Journal of Human Genetics, 28:165–173. https://doi:10.1038/s41431-019-0508-0

Ortiz-Quiroga, D., Ariza-Araújo, Y., & Pachajoa, H. (2018). Calidad de vida familiar en pacientes con síndrome de Morquio tipo IV-A. Una mirada desde el contexto colombiano (Suramérica). Rehabilitación, 52(4), 230–237. https://doi.org/10.1016/j.rh.2018.07.002

Oliveira, A. C. B., Paiva, S. M., Campos, M. R., & Czeresnia, D. (2008). Factors associated with malocclusions in children and adolescents with Down syndrome. American Journal of Orthodontics and Dentofacial Orthopedics, 133(4), 489–e1.

Pachajoa, H., Acosta, M. A., Alméciga-Díaz, C. J., Ariza, Y., Diaz-Ordoñez, L., Caicedo-Herrera, G., Cuartas, D., Nastasi-Catanese, J. A., Ramírez-Montaño, D., Silva, Y. K., Moreno, L., Satizabal, J., Garcia, N., Montoya, J., Prada, C., Porras, G., Velasco, H., & Candelo, E. (2021). Molecular characterization of mucopolysaccharidosis type IVA patients in the Andean region of Colombia. American Journal of Medical Genetics. Part C, Seminars in Medical Genetics, 187(3), 388–395. https://doi.org/10.1002/ajmg.c.31936

Park, S., Kim, J., Song, T.-Y., & Jang, D.-H. (2022). Case Report: The success of face analysis technology in extremely rare genetic diseases in Korea: Tatton–Brown–Rahman syndrome and Say–Barber –Biesecker–Young–Simpson variant of ohdo syndrome. Frontiers in Genetics, 13, 903199. https://doi.org/10.3389/fgene.2022.903199

Pascolini, G., Calvani, M., & Grammatico, P. (2022). First Italian experience using the automated craniofacial gestalt analysis on a cohort of pediatric patients with multiple anomaly syndromes. Italian Journal of Pediatrics, 48(1), 91. https://doi.org/10.1186/s13052-022-01283-w

Patterson D. (2009). Molecular genetic analysis of Down syndrome. Human Genetics, 126(1), 195–214. https://doi.org/10.1007/s00439-009-0696-8

Qiao, L., Yang, Y., Fu, P., Hu, S., Zhou, H., Peng, S., Tan, J., Lu, Y., Lou, H., Lu, D., Wu, S., Guo, J., Jin, L., Guan, Y., Wang, S., Xu, S., & Tang, K. (2018). Genome-wide variants of Eurasian facial shape differentiation and a prospective model of DNA based face prediction. Journal of Genetics and Genomics, 45(8), 419–432. https://doi.org/10.1016/j.jgg.2018.07.009

Quinto-Sánchez, M., Adhikari, K., Acuña-Alonzo, V., Cintas, C., Silva de Cerqueira, C.C., Ramallo, V., Castillo, L., Farrera, A., Jaramillo, C., Arias, W., Fuentes, M., Everardo, P., de Avila, F., Gomez-Valdés, J., Hünemeier, T., Gibbon, S., Gallo, C., Poletti, G., Rosique, J., Bortolini, M.C., Canizales-Quinteros, S., Rothhammer, F., Bedoya, G., Ruiz-Linares, A., & González-José, R. (2015). Facial asymmetry and genetic ancestry in Latin American admixed populations. American Journal of Physical Anthropology, 157:58–70. https://doi:10.1002/ajpa.22688

Reardon, W.; & Donnai, D. (2007). Dysmorphology demystified. Arch. Dis. Child. Fetal Neonatal Ed., 92, F225–F229.

Ruiz-Linares, A., Adhikari, K., Acuña-Alonzo, V., Quinto-Sanchez, M., Jaramillo, C., Arias, W., Fuentes, M., Pizarro, M., Everardo, P., de Avila, F., Gómez-Valdés, J., León-Mimila, P., Hunemeier, T., Ramallo, V., de Cerqueira, C.C.S., Burley, M-W., Konca, E., de Oliveira, M.Z., Veronez, M.R., Rubio-Codina, M., Attanasio, O., Gibbon, S., Ray, N., Gallo, C., Poletti, G., Rosique, J., Schuler-Faccini, L., Salzano, F.M., Bortolini, M-C., Canizales-Quinteros, S., Rothhammer, F., Bedoya, G., Balding, D., & Gonzalez-José, R. (2014). Admixture in Latin America: Geographic Structure, Phenotypic Diversity and Self-Perception of Ancestry Based on 7,342 Individuals. PLOS Genetics, 10: e1004572. https://doi:10.1371/journal.pgen.1004572

Rohlf, F. J., & Slice, D. (1990). Extensions of the Procrustes method for the optimal superimposition of landmarks. Systematic Biology, 39, 40–59.

Roper, R., & Reeves, R. (2006). Understanding the Basis for Down Syndrome Phenotypes. PLoS Genetics. 2. e50. 10.1371/journal.pgen.0020050.

Salzano, F.M., & Bortolini, M.C. (2002). The Evolution and Genetics of Latin American Populations. Cambridge University Press; Cambridge, p. 512.

Salzano, F. M., & Sans, M. (2014). Interethnic admixture and the evolution of Latin American populations. Genetics and Molecular Biology, 37(1 Suppl), 151–170. https://doi.org/10.1590/s1415-47572014000200003

Sawamoto, K., Álvarez González, J., Piechnik, M., Otero, F., Couce, M., Suzuki, Y., & Tomatsu, S. (2020). Mucopolysaccharidosis IVA: Diagnosis, Treatment, and Management. International Journal of Molecular Sciences, 21(4), 1517. https://doi.org/10.3390/ijms21041517

Schieppati, A., Henter, J-I., Daina, E., & Aperia, A. (2008). Why rare diseases are an important medical and social issue. The Lancet, 371:2039–2041. https://doi:10.1016/S0140-6736(08)60872-7

Sheehan, M.J., & Nachman, M.W. (2014). Morphological and population genomic evidence that human faces have evolved to signal individual identity. Nature Communications, 5:4800. https://doi:10.1038/ncomms5800

Starbuck, J. M., Cole, T. M., Reeves, R. H., & Richtsmeier, J. T. (2013). Trisomy 21 and facial developmental instability. American Journal of Physical Anthropology, 151(1), 49–57. https://doi.org/10.1002/AJPA.22255

Starbuck, J. M., Llambrich, S., Gonzàlez, R., Albaigès, J., Sarlé, A., Wouters, J., González, A., Sevillano, X., Sharpe, J., De La Torre, R., Dierssen, M., Vande Velde, G., & Martínez-Abadías, N. (2021). Green tea extracts containing epigallocatechin-3-gallate modulate facial development in Down syndrome. Scientific Reports, 11(1), 4715. https://doi.org/10.1038/s41598-021-83757-1

Stull, K. E., Tise, M. L., Ali, Z., & Fowler, D. R. (2014). Accuracy and reliability of measurements obtained from computed tomography 3D volume rendered images. Forensic Science International, 238, 133–140. https://doi.org/10.1016/j.forsciint.2014.03.005

Suárez-Guerrero, J. L., Suárez, A. K. B., Santos, M. C. V., & Contreras-García, G. A. (2013). Caracterización clínica, estudios genéticos, y manejo de la Mucopolisacaridosis tipo IV A. Medicas UIS, 26 (2):43–50.

Suárez-Guerrero, J. L., Gómez Higuera, P. J. I., Arias Flórez, J. S., & Contreras-García, G. A. (2016). Mucopolisacaridosis: Características clínicas, diagnóstico y de manejo. Revista Chilena de Pediatría, 87(4), 295–304. https://doi.org/10.1016/j.rchipe.2015.10.004

Suárez-Obando, F. (2018). La atención clínica de las enfermedades raras: un reto para la educación médica. Medicina-Buenos Aires, 40, 228–241.

Terrazas, K., Dixon, J., Trainor, P.A. & Dixon, M.J. (2017). Rare syndromes of the head and face: mandibulofacial and acrofacial dysostoses. Wiley Interdisciplinary Reviews: Developmental Biology, 6: e263. https://doi-org.sire.ub.edu/10.1002/wdev.263

Urrea-Giraldo, F., & Álvarez, A. F. C. (2017). Cali, an enlarged region city: An approximation from the ethnic-racial dimension and population flows. Revista Sociedad y Economía UV, 33: 145–174. https://doi.org/10.25100/sye.v0i33.5628

Valencia Arana, C. A., González Arango, R., Gaitán Quintero, L. M., Naranjo Cardona, A. L., Giraldo Ospina, G. A., & Castaño Molina, E. (2008). Prevalencia al nacimiento de síndrome de Down en la ciudad de Manizales (Caldas-Colombia) durante el periodo 2004-2005. Biosalud, 69.

Visnapuu, V., Peltonen, S., Alivuotila, L., Happonen, R.-P., & Peltonen, J. (2018). Craniofacial and oral alterations in patients with Neurofibromatosis 1. Orphanet Journal of Rare Diseases, 13(1), 131. https://doi.org/10.1186/s13023-018-0881-8

Viteri, J., Carrasco, A. M., Jácome, M., Vaca, G., Tubón, I., Rodríguez, V., Morales, M. F., & Vinueza, D. (2020). Enfermedades huérfanas. Archivos Venezolanos de Farmacología y Terapéutica, 39 (5), 627–636.

Xiong, Z., Dankova, G., Howe, L.J., Lee, M.K., Hysi, P.G., de Jong, M.A., Zhu, G., Adhikari, K., Li, D., Li, Y., Pan, B., Feingold, E., Marazita, M.L., Shaffer, J.R., McAloney, K., Xu, S-H., Jin, L., Wang, S., de Vrij, F.M., Lendemeijer, B., Richmond, S., Zhurov, A., Lewis, S., Sharp, G.C., Paternoster, L., Thompson, H., Gonzalez-Jose, R., Bortolini, M.C., Canizales-Quinteros, S., Gallo, C., Poletti, G., Bedoya, G., Rothhammer, F., Uitterlinden, A.G., Ikram, M.A., Wolvius, E., Kushner, S.A., Nijsten, T.E., Palstra, R-JT., Boehringer, S., Medland, S.E., Tang, K., Ruiz-Linares, A., Martin, N.G., Spector, T.D., Stergiakouli, E., Weinberg, S.M., Liu, F., & Kayser, M. (2019). Novel genetic loci affecting facial shape variation in humans. eLife, 8: e49898. https://doi:10.7554/eLife.49898

